# Perspective on Harnessing Large Language Models to Uncover Insights in Diabetes Wearable Data

**DOI:** 10.1101/2024.07.29.24310315

**Authors:** Arash Alavi, Kexin Cha, Delara P Esfarjani, Bhavesh Patel, Jennifer Li Pook Than, Aaron Y. Lee, Camille Nebeker, Michael Snyder, Amir Bahmani

## Abstract

Large Language Models (LLMs) have gained significant attention and are increasingly used by researchers. Concurrently, publicly accessible datasets containing individual-level health information are becoming more available. Some of these datasets, such as the recently released Artificial Intelligence Ready and Equitable Atlas for Diabetes Insights (AI-READI) dataset, include individual-level data from digital wearable technologies. The application of LLMs to gain insights about health from wearable sensor data specific to diabetes is underexplored. This study presents a comprehensive evaluation of multiple LLMs, including GPT-3.5, GPT-4, GPT-4o, Gemini, Gemini 1.5 Pro, and Claude 3 Sonnet, on various diabetes research tasks using diverse prompting methods to evaluate their performance and gain new insights into diabetes and glucose dysregulation. Notably, GPT-4o showed promising performance across tasks with a chain-of-thought prompt design (aggregate performance score of 95.5%). Moreover, using this model, we identified new insights from the dataset, such as the heightened sensitivity to stress among diabetic participants during glucose level fluctuations, which underscores the complex interplay between metabolic and psychological factors. These results demonstrate that LLMs can enhance the pace of discovery and also enable automated interpretation of data for users of wearable devices, including both the research team and the individual wearing the device. Meanwhile, we also emphasize the critical limitations, such as privacy and ethical risks and dataset biases, that must be resolved for real-world application in diabetes health settings. This study highlights the potential and challenges of integrating LLMs into diabetes research and, more broadly, wearables, paving the way for future healthcare advancements, particularly in disadvantaged communities.

## Introduction

Large language models (LLMs) like GPT^1^, Gemini^2^, and Claude^3^ are being widely adopted due to their exceptional abilities in natural language processing, data analysis, and generating insightful and coherent text, enabling more efficient and accurate analysis of complex datasets across various domains^4,5^. These models are frequently updated based on user data, feedback, and design improvements, enhancing their capabilities over time. Their remarkable potential to adapt to various domains and uncover new insights, which can be more facile and less time-consuming than traditional artificial intelligence (AI) and machine learning (ML) methods, makes them particularly valuable. The application of LLMs in healthcare has garnered significant interest from researchers^6–8^. However, there is still a gap between the potential capabilities of LLMs and their application to real-time physiological data gathered from wearables. This is a rapidly growing domain, with an increasing number of users (both researchers and the end user wearing the watch) employing smartwatches to track vital signs such as heart rate, steps, and respiratory rate, as well as glucose monitoring devices to track blood sugar levels^9–12^.

To bridge this gap and demonstrate practical and equitable real-world applications, we conducted a study to evaluate whether state-of-the-art LLMs can quickly and effectively answer research questions and uncover new insights from wearable data, specifically focusing on blood glucose levels, collected from a moderate number of participants. Our focus is primarily on diabetes-related data, but we also extend our analysis to other physiological metrics such as heart rate and respiratory rate gathered from smartwatches.

For this study, we utilized the AI-READI (Artificial Intelligence Ready and Equitable Atlas for Diabetes Insights) dataset^13^. We leveraged cutting-edge LLMs (i.e. GPT^1^, Gemini^2^, and Claude^3^) and applied various prompting methods to different diabetes-related health tasks. We compared the performance of these LLMs using multiple prompting techniques and evaluation metrics. Finally, we used the best-performing model to extract new insights from the diabetes dataset, uncovering information that might be overlooked by traditional AI/ML methods or would require a time-consuming process. This study aims to bridge the gap and showcase the practical benefits of LLMs in utilizing wearable data to gain valuable health insights, with a specific focus on diabetes.

### LLMs in healthcare research

In general healthcare, LLMs like GPT-4 are enhancing various medical practices by improving diagnostic accuracy, streamlining administrative tasks, and supporting clinical decision-making^14,18^. These models can analyze and interpret vast amounts of medical data, including patient records, radiology reports, and clinical notes, thus aiding healthcare professionals in making more informed decisions. For instance, LLMs have been used to generate accurate patient discharge summaries, draft clinical notes, and even assist in medical education by providing up-to-date medical information and recommendations^14–18^. Despite their potential, there are challenges such as ensuring data privacy, addressing biases in training data, and maintaining the accuracy and reliability of the outputs^19,20^.

In precision medicine, LLMs are playing a crucial role by enabling personalized treatment plans based on individual patient data. These models can analyze genetic information, lifestyle factors, and environmental influences to predict disease risks and treatment responses. For example, LLMs have been employed to identify appropriate treatments for cancer patients, support decision-making in tumor boards, and predict disease progression in neurodegenerative disorders^21,22^. The ability of LLMs to integrate and analyze multimodal data—such as genomic sequences, imaging data, and electronic health records—makes them invaluable in tailoring treatments to individual patients, thereby improving outcomes and reducing adverse effects^23–25^.

Wearables represent a potential significant application area for LLMs in healthcare. These devices generate continuous streams of physiological and longitudinal data, which can be analyzed in real-time to monitor patient health, detect anomalies, and predict potential health issues. LLMs can process the data from wearables to provide insights into a patient’s daily health metrics, offer personalized health recommendations, and even predict the onset of diseases like cardiovascular conditions or diabetes^11,26–29^. The integration of LLMs with wearable technology facilitates proactive health management, enabling real-time delivery of care and health insights without requiring patients to directly interface with a physician or actively engage in a mobile health app.

### Approach

We wished to evaluate different LMMs for their ability to accurately address tasks as well as gain new insights into glucose dysregulation. The tasks we chose were of increasing order of complexity. Our overall approach to utilize prompt-driven techniques for enhanced diabetes insights is as follows: We first describe the dataset and the data preprocessing and integration steps utilized. Next, we detail the LLMs employed in our experiment. We then present our experimental design, which involves utilizing LLMs to address multiple diabetes insight tasks through various prompting strategies, including zero-shot prompting, few-shot prompting, and chain-of-thought prompting. Our approach to prompt engineering remains model-agnostic throughout these steps.

We utilized the dataset from the AI-READI project, an AI-ready and ethically-sourced dataset aimed at supporting research related to type 2 diabetes (NIH Bridge2AI program). The public dataset includes health information, encompassing diabetes status, blood sugar levels gathered from the Dexcom G6 Continuous Glucose Monitoring (CGM) device -- 197 participants --, and fitness tracker data (e.g., heart rate, stress, respiratory rate, activity) gathered from a Garmin smartwatch -- 135 participants. This data is representative of a diverse group of participants, balanced across sexes (equal numbers of male and female), ethnicities/races (equal numbers of Asian, Black, Hispanic, and White), and health states (equal numbers of non-diabetic, diet-controlled diabetic, oral medication-controlled diabetic, and insulin-controlled diabetic). We then synchronized and aggregated the data and ensured that all participants under analysis had sufficient numbers of blood sugar level measurements and wearable data, with the criterion being at least one day of data for all data types. The resulting synchronized subset includes data for a total of 113 participants, comprising 42 diabetic and 71 non-diabetic individuals.

For the LLMs, we utilized the most commonly and widely used models to conduct our analysis and experiments, including **GPT-3.5**^30^, **GPT-4**^31^, **GPT-4o**^32^, **Gemini**^33^, **Gemini 1.5 pro**^34^, and **Claude 3 Sonnet**^35^. These LLMs were selected due to their widespread adoption and proven effectiveness in various natural language processing applications. Studies have evaluated the performance of these LLMs from various aspects, including conversational QA tasks^36^, logical reasoning^37,38^, coding tasks^37^, understanding electronic health records^39^, and ethical perspectives^40^. These analyses reveal the strengths and limitations of each LLM. Here, by incorporating these models, we aimed to leverage their advanced capabilities and robust performance to ensure a comprehensive evaluation across different state-of-the-art LLMs.

We chose tasks of increasing order of complexity using different prompting approaches. **Fig. 1** illustrates the prompt design utilized in this study beginning with **zero-shot prompting**. This approach involves crafting a straightforward prompt that directly addresses the diabetes-related research question without incorporating additional domain-specific or engineering context, which can be beneficial in maintaining clarity and focus. Following this, we employed **few-shot prompting**, which introduces some domain-specific information by providing examples (labels) that enable the model to learn relevant knowledge and thereby improve its performance. Finally, we utilized **chain-of-thought prompting**, where the LLM is presented with examples of chained, step-by-step logical reasoning (via additional health context or user context). This method aids the model in developing a more structured and coherent approach to solving the problem, ultimately leading to better solutions.

**Figure 1.**
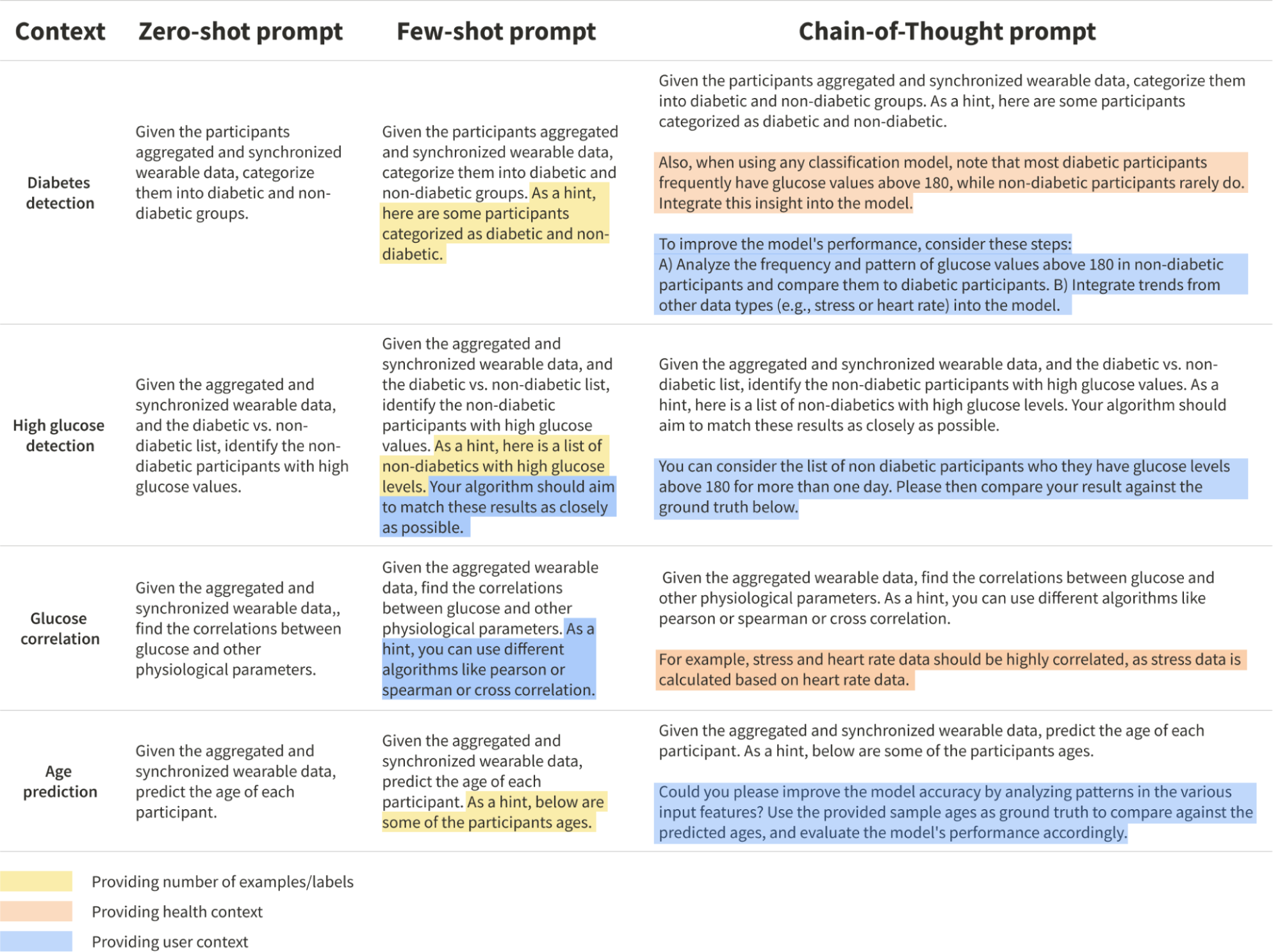
Different types of contexts and three prompting settings (zero-shot, few-shot, and chain-of-thought).

As shown in **Fig. 1**, we categorized our questions into four distinct tasks. The first task involves querying the LLM to predict diabetes by analyzing participants’ glucose level and other wearable data (**Diabetes detection**). The second task focuses on identifying non-diabetic participants who still experience periods of high glucose levels (**High glucose detection**). The third task examines the correlation between glucose levels and other types of wearable data, aiming to uncover any significant relationships between different types of physiological data (**Glucose correlation**). Lastly, the fourth task aims to predict participants’ ages based on their glucose levels and other wearable data (**Age prediction)**. This structured approach allows us to comprehensively explore various aspects of the dataset and evaluate the LLMs’ capabilities for different types of analysis.

### LLMs performance evaluation

For the first and second tasks, we use the F1-score as a balanced measure of precision and recall to compare the performance of the LLMs. For the third task, we evaluate the models’ ability to correctly identify three types of correlations between glucose and wearable data: Pearson, Spearman, and Cross correlation. This assessment helps us understand the models’ proficiency in capturing different correlations within the data. For the fourth task, we use the Mean Absolute Error (MAE) to compare the performance of the models in predicting participants’ ages. Additionally, we apply min-max normalization to the MAE values to ensure comparability and consistency in the evaluation metrics across different tasks.

**Fig. 2** summarizes the performance among the six models tested with a focus on diabetes. As illustrated, among all models, GPT-4o exhibits the best performance when using the chain-of-thought prompting method. The next best model is Gemini 1.5 pro, which, interestingly, excels with the few-shot prompting approach. This is particularly evident in the age prediction task in this model, where providing direct examples of participants’ ages makes it easier for the model to perform the task, compared to incorporating additional reasoning features, such as other wearable data (e.g., heart rate, respiratory rate, and stress level), into the process. This finding highlights the importance of selecting the appropriate prompting method for different models and tasks to achieve optimal performance. Another important observation is that GPT-4o performs better than GPT-4 and significantly better than GPT-3.5, reflecting the advancements in natural language processing, contextual comprehension, and response generation capabilities in these successive model iterations. This supports previous research, which shows that the behavior of the ‘same’ LLM service can change substantially in a relatively short period via updates to the models^41^.

**Figure 2.**
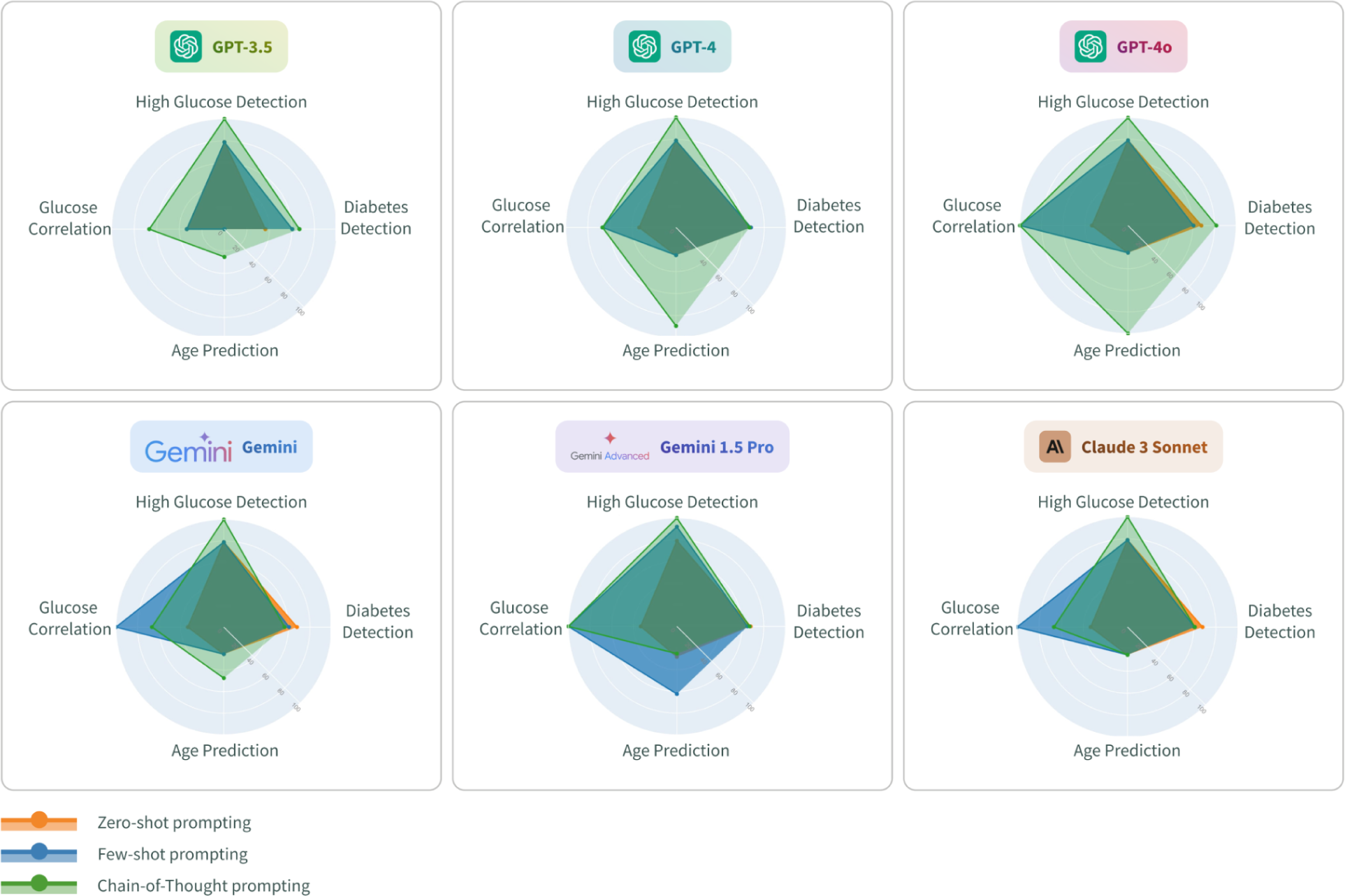
Overview of LLMs performance under three prompting settings. For the various tasks, we employ the following metrics: for “diabetes detection” and “high glucose detection,” we use the F1-score; for the “glucose correlation” task, we utilize three types of correlations: Pearson, Spearman, and cross-correlation; and for the “age prediction” task, we use the Mean Absolute Error (MAE). The aggregate performance score for all LLMs, calculated using the optimal prompting settings for each model is as follows: **GPT-3.5: 64.6%**, **GPT-4: 80.7%**, **GPT-4o: 95.5%**, **Gemini: 67.7%**, **Gemini 1.5 pro: 79.9%**, and **Claude 3 Sonnet: 66.2%.**

### Diabetes insights using LLMs

An LLM-based module can be immensely helpful for researchers in discovering novel insights at various stages of their analysis. Initially, it can serve as a powerful tool for generating hypotheses by identifying trends and anomalies in large datasets. During intermediate stages, it can assist in refining these hypotheses by providing contextual understanding and suggesting relevant variables to consider. Finally, as part of the final analysis, an LLM can validate findings and offer new perspectives, ensuring a comprehensive exploration of the data. It is also important to note that these tasks are unlikely to have been part of the training corpus for these LLMs, as they are relatively new and specialized biomedical tasks, and it is remarkable that the LLMs demonstrated any capability to perform these tasks. Hence, this capability of LLMs to assist throughout the research process not only enhances the efficiency and depth of analysis but also fosters innovative discoveries that might otherwise be overlooked.

Since GPT-4o (using chain-of-thought prompting) demonstrated the best performance on our proposed diabetes-based research prompts, we used this model to investigate how this LLM can uncover new insights in the AI-READI dataset. Leveraging its advanced capabilities, GPT-4o was able to identify patterns and correlations that were not immediately obvious, providing a deeper understanding of the data and potentially highlighting areas for further study.

First, we started with analyzing the glucose level distributions in diabetic versus non-diabetic participants. **Fig. 3A** displays the glucose level distribution for 42 diabetic and 71 non-diabetic participants. As expected, the majority of diabetic participants frequently experience high glucose levels (over 180). However, some non-diabetic participants also have significant periods of high glucose, as identified through prompting over task two in the previous section. Next, we utilized GPT-4o to explore the correlations between glucose and stress, and glucose and respiratory rate, in both diabetic and non-diabetic participants. These findings are illustrated in **Fig. 3B** and **3C**, respectively. **Fig. 3B** reveals that changes in glucose levels (from normal to high) are significantly associated with stress levels in diabetic participants compared to non-diabetic participants as diabetic participants are more likely to experience high stress levels during periods of elevated glucose. Regarding respiratory rate, as shown in **Fig. 3C**, the majority of both diabetic and non-diabetic participants maintained a normal respiratory rate during periods of normal and high glucose levels. This indicates that glucose level changes in both groups do not significantly affect respiratory rate. However, diabetic participants still exhibited more frequent episodes of tachypnea. One explanation for the lack of significant differences in respiratory rates between diabetic and non-diabetic participants in this dataset is the insufficient duration of aggregated data in the AI-READI dataset, which encompasses a maximum of 11 days of data across all participants. This analysis provides a deeper understanding of how these variables interact differently across the two groups and provide new insights into the management and prediction of glucose levels. Interestingly, stress tracking relies on heart rate (HR) and heart rate variability (HRV) data. These findings from the LLM corroborate their biological associations to higher glucose levels in the diabetic population^42^.

**Figure 3.**
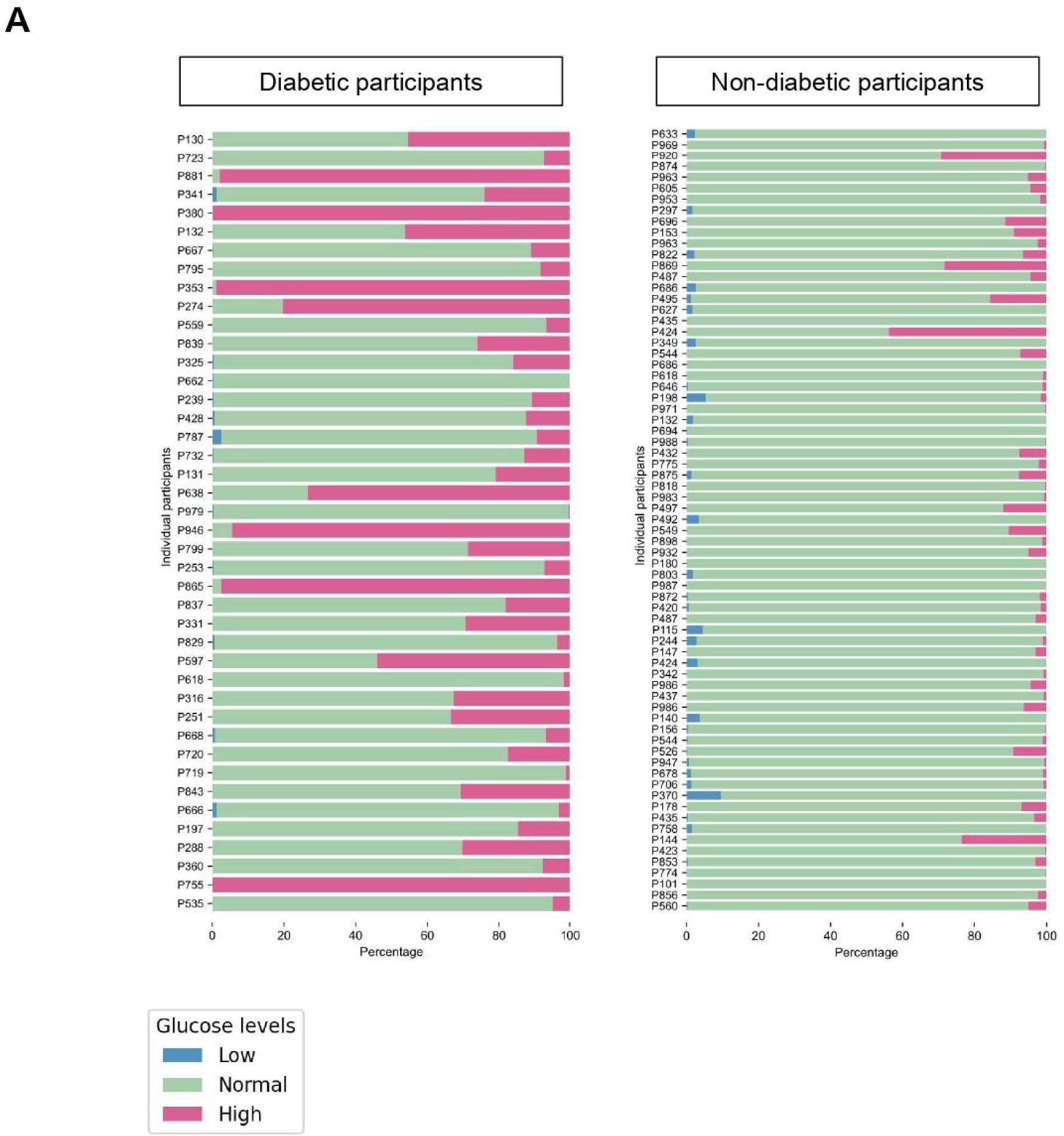
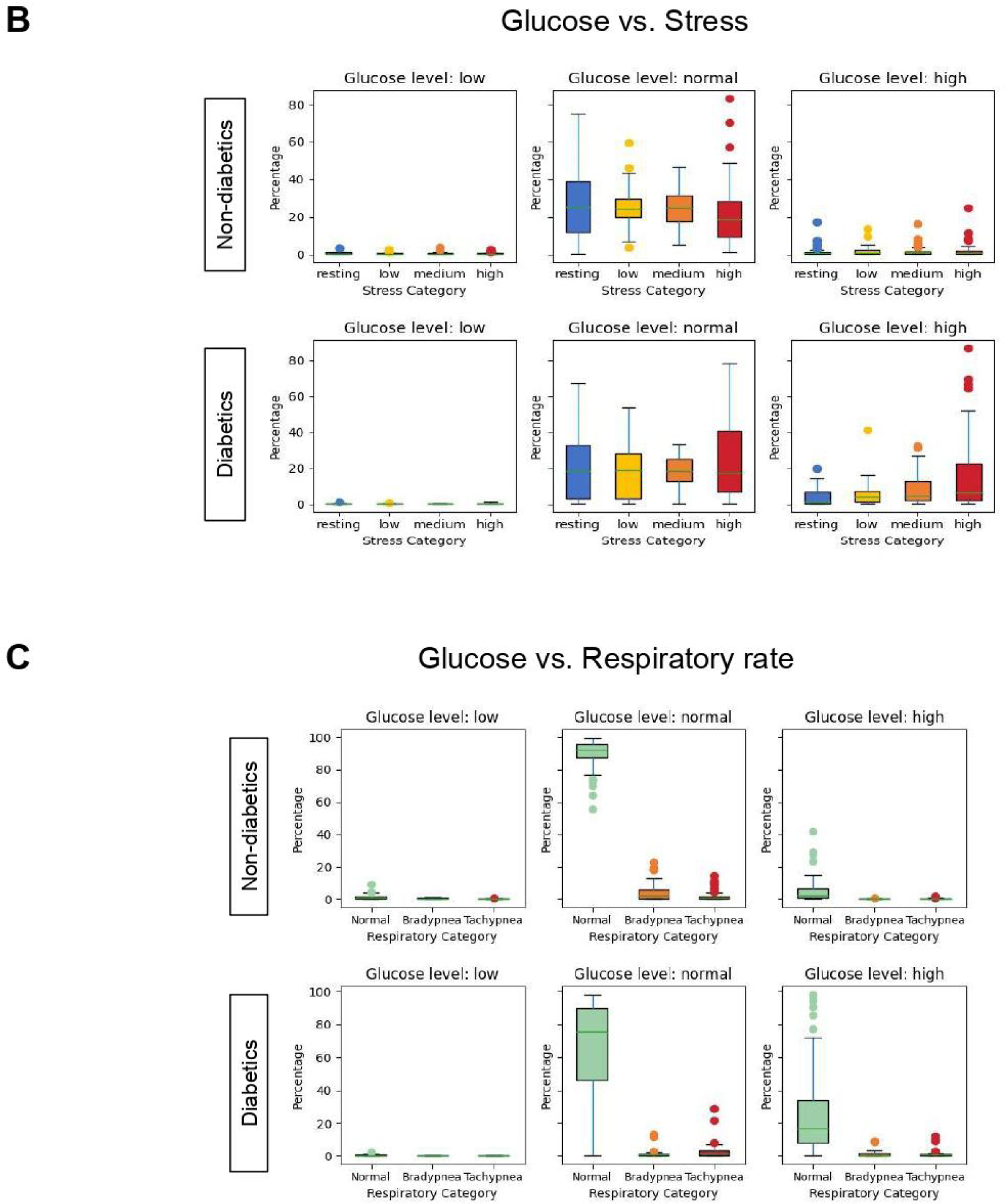
Diabetes Insights. **(A)** Glucose level distribution for diabetic vs. non-diabetic participants. **(B)** Glucose vs. stress in diabetic and non-diabetic participants with respect to three categories of glucose level (low, normal, and high) and four categories of stress (resting, low, medium, and high). **(C)** Glucose vs. respiratory rate in diabetic and non-diabetic participants with respect to three categories of glucose level (low, normal, and high) and three categories of respiratory rate (normal, bradypnea, and tachypnea).

### Knowledge gaps and potential limitations

While we demonstrated the practical application of LLMs in uncovering new insights in glucose and wearable data, this approach does have a few limitations: **Model understanding:** LLMs may struggle to interpret raw wearable data accurately without proper preprocessing and contextual information. This is why we apply data preprocessing and synchronization before running LLMs on the dataset^43,44^. **Computational resources:** Processing large volumes of wearable data and running complex analyses via LLMs can require significant computational power and memory, sometimes exceeding the LLM’s token limitations^45^. **Ethical, legal and social implications:** Handling sensitive health data via LLMs raises concerns about data privacy and security. Ensuring compliance with regulations such as HIPAA is crucial. Moreover, wearable data and analyses may reflect demographic biases, leading to biased insights that LLMs might overlook^46,47^. In this study, we used the paid version of the LLMs with data privacy agreements that ensured our dataset content would not be used to train their models, as approved by the LLM providers. Additionally, we did not include race, ethnicity, and other demographic information in our experiments. Future work needs to consider the diversity and equitability of the training datasets used in LLMs. Although there is increasing awareness of the importance of inclusion and the use of diverse training datasets in LLM model learning, including additional reinforcement learning with human interface^48^, further efforts are necessary to ensure checks and balances are in place to reduce bias and mitigate disparities. **Validation of insights:** Insights generated by LLMs need to be rigorously validated through clinical trials and studies to ensure their accuracy and efficacy^49–51^.

## Conclusion

While LLMs have garnered significant attention across various research domains, there has yet to be a comprehensive study demonstrating their potential to revolutionize diabetes research by offering new insights from wearable data. This paper presents the first proof of concept, illustrating the benefits of utilizing LLMs to analyze and derive new insights from publicly available wearable data, with a focus on diabetes. Our findings highlight that GPT-4o, the latest and most advanced model from OpenAI, outperforms others, though models like Gemini 1.5 Pro also show promising results. Future research is essential to address existing limitations, including computational resource demands, ethical and privacy concerns, the need for more equitable data representation, and rigorous medical validation of new findings. Our findings highlight the need for cost-effective healthcare solutions, such as using wearable technologies to monitor basic physiological metrics like heart rate and glucose levels. These accessible methods are essential for reducing financial burdens on economically disadvantaged and historically marginalized communities. Additionally, integrating LLMs can enhance data analysis without requiring expensive infrastructure. While advanced solutions have their place, their higher costs and complexity may disadvantage vulnerable groups. Therefore, we advocate for a healthcare model focused on affordability, accessibility, and strategic technology use to ensure equitable health outcomes.

## Data Availability

Source data can be found from the following link: https://doi.org/10.60775/fairhub.1.

## Code availability

All code for different LLMs and prompting settings used in the manuscript are publicly available under the MIT license at: https://github.com/StanfordBioinformatics/llm-diabetes.

## Acknowledgments

This work was supported by NIH grant OT2OD032644 and departmental funding from the Stanford Genetics department. We acknowledge Amazon Web Services, OpenAI, and Stanford Genetics Bioinformatics Service Center (GBSC) for this research.

## Contributions

All authors read, revised and approved the manuscript.

